# Are high urea values before intravenous immunoglobulin replacement a risk factor for COVID-related mortality?

**DOI:** 10.1101/2022.01.29.22270080

**Authors:** Gökhan Aytekіn, Emel Atayik

**Author notes:** **Corresponding Author:** Gokhan AYTEKIN, MD, University of Health Sciences, Konya City Hospital, Division of Allergy and Clinical Immunology, 42080, Konya Turkey, **Email:**. **Informed consent** The study protocol was approved by the Ethics committee of the Karatay University (with the decision dated 09.02.2021, decision number: 2020/021). Informed consent was obtained from study participants. Manuscripts that have not been presented orally or as a poster. **Funding source:** No funding was secured for the study. **Financial disclosure:** The authors have no financial relationship relevant to this article to disclose. **Disclaimers:** None.

## Abstract

**Objective:** **S**ince the World Health Organization accepted The Coronavirus Disease 2019 (COVID-19) as a pandemic and there is still no effective treatment, it becomes crucial that the physicians interested in COVID-19 treatment share all the data they acquire, particularly in vulnerable patient groups, to reduce morbidity and mortality.

**Methods:** The study included 81 adult (Female: 27, Male: 54) COVID-19 patients who were hospitalized for the treatment of COVID-19 between April 2020 and September 2020 and were followed-up, treated and consulted in the immunology clinic for intravenous immunoglobulin (IVIG) treatment.

**Results:** The univariate analysis found that the number of days of hospitalization in service, being intubated, number of IVIG treatment days, and the urea value before IVIG treatment were independent risk factors for mortality (p:0.043, p:0.001, p:0.074, p:0.004, respectively). As a result of multivariate analysis, being intubated and urea value before IVIG treatment were found to be independent risk factors for mortality (p:0.001 and p:0.009).

It was found that for 60 mg/dL level of urea value before IVIG treatment, the sensitivity value for mortality in COVID-19 patients was 46.2%, and the specificity was 35.5% (p:0.029)

**Conclusion:** The study found that urea values before IVIG treatment were a risk factor for mortality in patients who received IVIG treatment for COVID-19. This is important as it indicates that BUN values should be closely monitored in patients given IVIG treatment for COVID-19. It also suggests that when resources are limited and risk stratification is required in COVID-19 patients, BUN values can be helpful.

## 1. Introduction

The coronavirus disease 2019 (COVID-19), caused by severe acute respiratory syndrome coronavirus-2 (Sars-CoV-2), has affected the whole world in economic, social, spiritual, and many other areas, particularly in the field of health, since December 2019, when it was first described (1, 2). As the disease is highly contagious, the virus spread worldwide in a short time and caused one of the most catastrophic pandemics in human history (3). Although there are some vaccines to reduce virus transmission and develop protection against it, it is obvious that vaccinating all the people in the world will not be possible in such short term. Although it has been more than one year since the WHO (World Health Organization) accepted COVID-19 as a pandemic, there is still no effective treatment. Until now, many treatment options, particularly antimalarial drugs and antivirals, systemic corticosteroids, tocilizumab, anakinra, conventional plasma therapy, and intravenous immunoglobulin (IVIG) therapy, have been tried in the form of monotherapy or combinations for treating COVID-19, there is still no consensus on its treatment (4-7).

For this reason, it becomes crucial that the physicians interested in COVID-19 treatment share all the data they acquire, particularly in vulnerable patient groups, to reduce morbidity and mortality. Regarding COVID-19 treatment management, many countries have created their treatment protocols, and many associations have published guidelines for its treatment. COVID-19 treatment in Turkey has been primarily applied in line with the TC Ministry of Health protocols. In general, the patients positive for SARS-CoV-2 Polymerase Chain Reaction (PCR) (+) were put on hydroxychloroquine and favipiravir treatment at appropriate doses. Patients who did not benefit from these treatments and/or had underlying risk factors were hospitalized. In addition to respiratory support treatments, patients were treated with conventional plasma, systemic steroid therapy, immunomodulatory therapies such as tocilizumab and anakinra, and IVIG treatment, whichever appropriate, as line therapies (8). IVIG was administered as per the clinical immunologists’ opinions and in the proper dose and time intervals.

Considering that pulmonary lesions in COVID-19 are caused by viral infiltrates and inflammatory response, IVIG treatment provides inflammatory cytokine balance, inhibits auto-reactive T cells, reduces antibody production from CD19^+^ B cells, and reduces macrophage activity. The IVIG treatment is thought to provide a regression in pulmonary lesions, reducing the need for mechanical ventilation, length of hospital stay, and mortality rates in these patients (9, 10). Therefore, this study aimed to retrospectively examine the data of patients who reported to the immunology clinic for IVIG treatment in a tertiary referral hospital and who were hospitalized, followed up, and treated for COVID-19. The study also investigated the effects of the patients’ clinical, laboratory, and treatment characteristics and risk factors for mortality in patients with COVID-19 treating with IVIG treatment.

## 2. Methods

### Study design and study population

The study included 81 adult (Female [F]: 27, 33.3%; Male [M]: 54, 66.7%) COVID-19 patients who were hospitalized for the treatment of COVID-19 in a tertiary referral hospital between April 2020 and September 2020 and were followed-up, treated and consulted in the immunology clinic for IVIG treatment. A review of medical records (including information on age, sex, disease duration) was undertaken. Venous blood samples for biochemical analyses were drawn after at least ten hours of fasting, early in the morning. All biochemical analyses were conducted in the Central Biochemistry Laboratory of the Konya Education and Research Hospital.

Complete blood counts were performed using Sysmex XN-10 (Sysmex Corporation, Kobe, Japan) analyzers with the fluorescent flow cytometry method. Serum creatinine levels were measured using the Jaffe method. Quantitative determination of serum IgG, IgM, IgA, and IgE was done through particle-enhanced immunonephelometry using the Siemens BN II/BN ProSpec system (Erlangen, Germany).

The follow-up period of all patients started with their hospitalization. For the patients who died, the number of days between the date of hospitalization and death was accepted as the follow-up period. The duration of follow-up was calculated by confirming whether the discharged patients were alive or not through the TC Death Reporting System 2 weeks after discharge. For patients who died within two weeks of discharge, the follow-up period was accepted as the number of days between the date of hospitalization and death. For patients who lived more than two weeks after discharge, the follow-up period was calculated by adding 14 days to the number of days they stayed in the hospital.

The time until hospitalization, resulting from the emergence of SARS-CoV-2-related symptoms such as fever, cough, and body pain, was considered the duration of illness. The duration of the follow-up in the service was specified as the day of hospitalization and the follow-up period in the intensive care unit as the duration of intensive care hospitalization. All patients in the study received IVIG treatment. Some patients were followed only in the service and received IVIG treatment in the service. Some patients received IVIG treatment in the intensive care unit. Patients who received IVIG treatment in the service and those who received IVIG treatment in the first 24/48 h after their admission to intensive care were specified as the IVIG treatment ICU first 24/48 h.

The systemic inflammatory index (SII) was calculated by the formula platelet x neutrophil/lymphocyte. The SARS-CoV-2 diagnosis was established with the detection of the SARS-CoV-2 genome via the PCR method from the nasopharyngeal sample (nasal swab) in patients with symptoms suggestive of SARS-CoV-2 infection such as fever, cough, shortness of breath, joint and body pain, and/or viral infiltration on lung imaging (PA chest radiography or lung tomography).

The permission for the study was obtained from the Republic of Turkey, Ministry of Health Scientific Research Platform. In addition, an ethics committee approval was obtained from Karatay University Ethics Committee (with the decision dated 09.02.2021, decision number: 2020/021). Written informed consent was obtained from each patient. The study was conducted as per the principles of the Declaration of Helsinki.

### Statistical Analyses

Statistical analyses were performed using the SPSS version 22.0 software package (IBM Corp., Armonk, NY, USA). Normally distributed parameters were presented as mean ±standard deviation, and data that were not normally distributed were expressed as median (interquartile range: minimum-maximum). Descriptive data were presented as frequencies and percentages and compared using the Chi-square test. Comparisons between baseline characteristics were performed by independent Student t, Mann-Whitney rank-sum, Fisher’s exact or Chi-square tests where appropriate. Independent predictors for mortality were determined using binomial logistic regression analysis, Cox regression analysis, and Kaplan-Meier test. ROC curves are used to choose the most appropriate cut-off for urea level. A *p*-value of <0.05 was considered statistically significant.

## 3. Results

A total of 81 patients, 27 of whom were women (33.3%), were included in the study. The average age of the patients was 71 (26–94) years. During the follow-up, the mortality rate was 64.2%. The rate of intubated patients was 45.7%. The average follow-up period was 19 (1–38) days. The duration of hospitalization was 17 (1–38) days, and the duration of hospitalization in intensive care was 10 (0–30) days. All patients received hydroxychloroquine and favipiravir treatment during their follow-up. In addition, IVIG treatment was given to all patients. While 35 patients (43.2%) received tocilizumab treatment, 15 patients (18.5%) received conventional plasma and 33 patients (40.7%) received pulse steroid treatment. 61.7% of the patients in the first 24 h of their admission to intensive care, and 64.2% of the patients in the first 48 h of their admission to intensive care received IVIG treatment. The demographic, laboratory, and clinical characteristics of the patients have been summarized in Table 1.

**Table 1:**
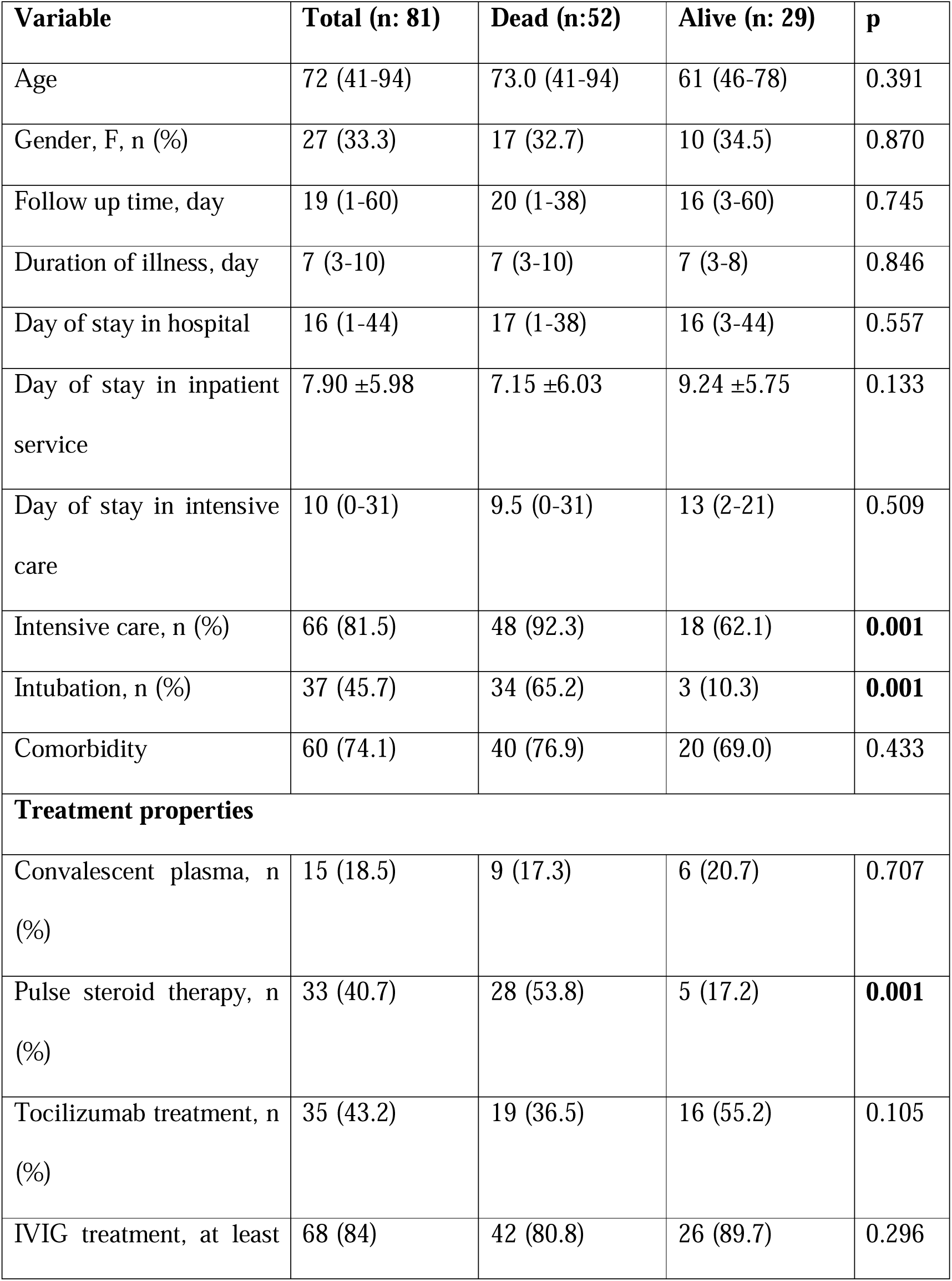
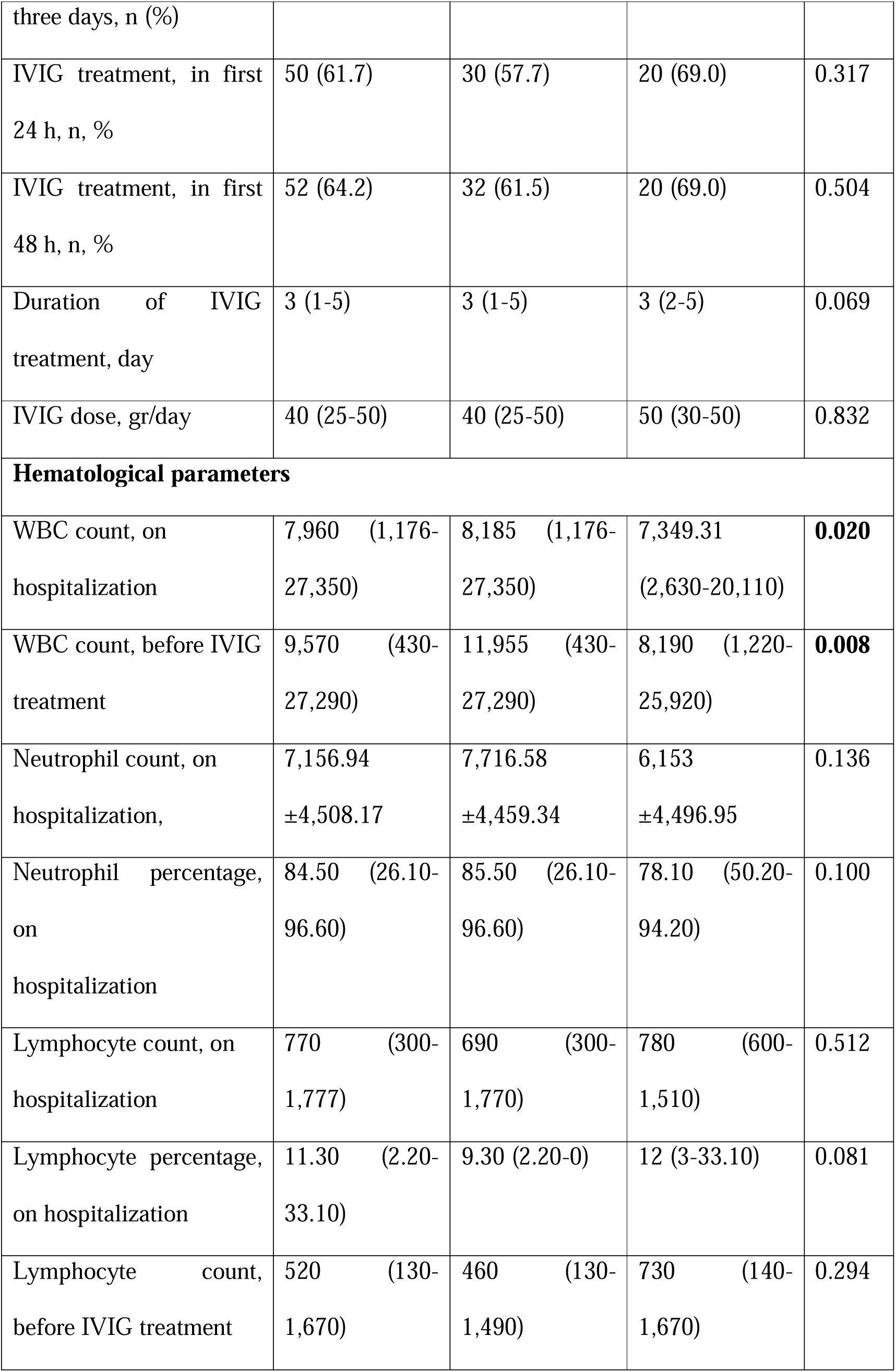
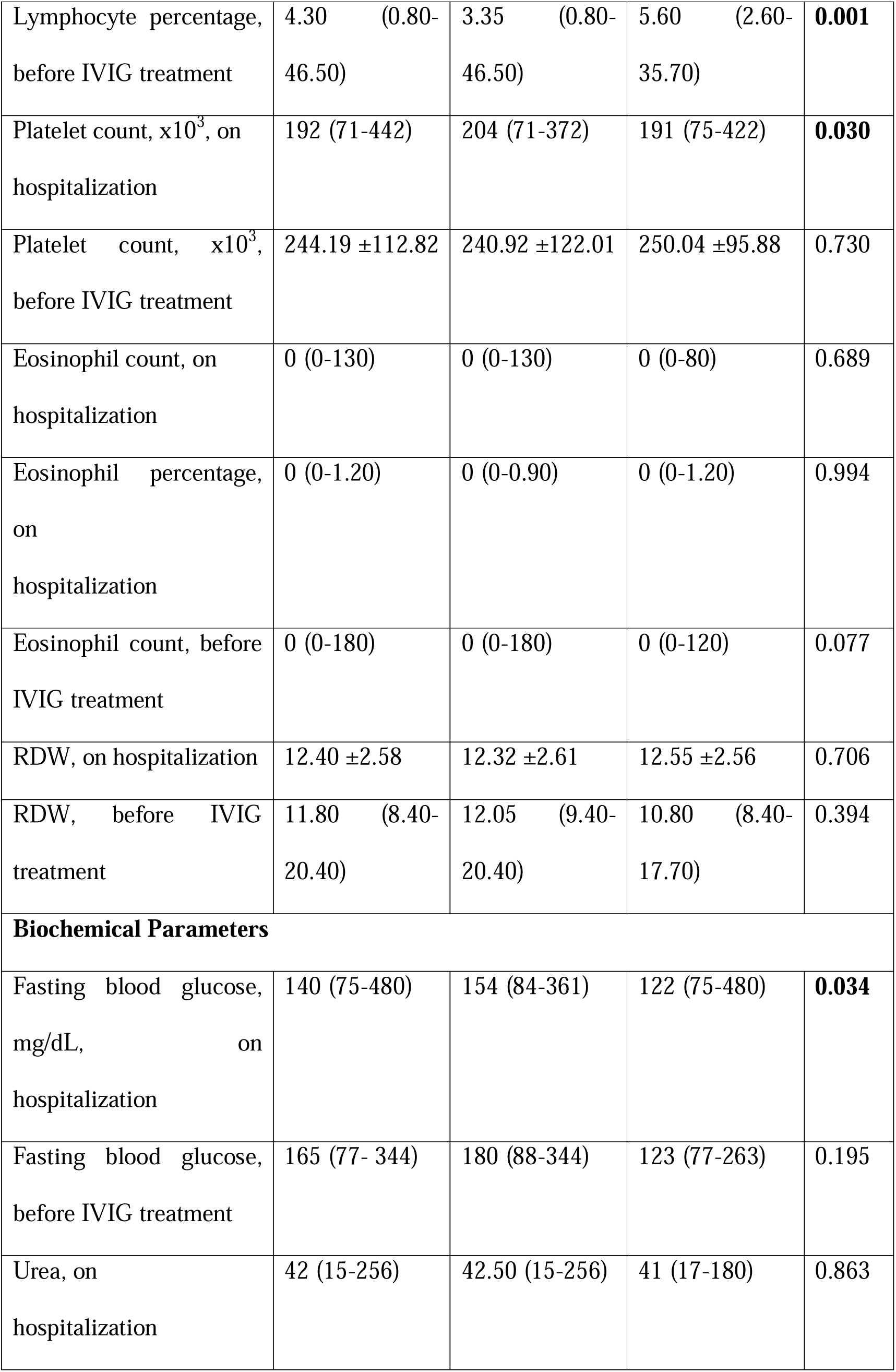
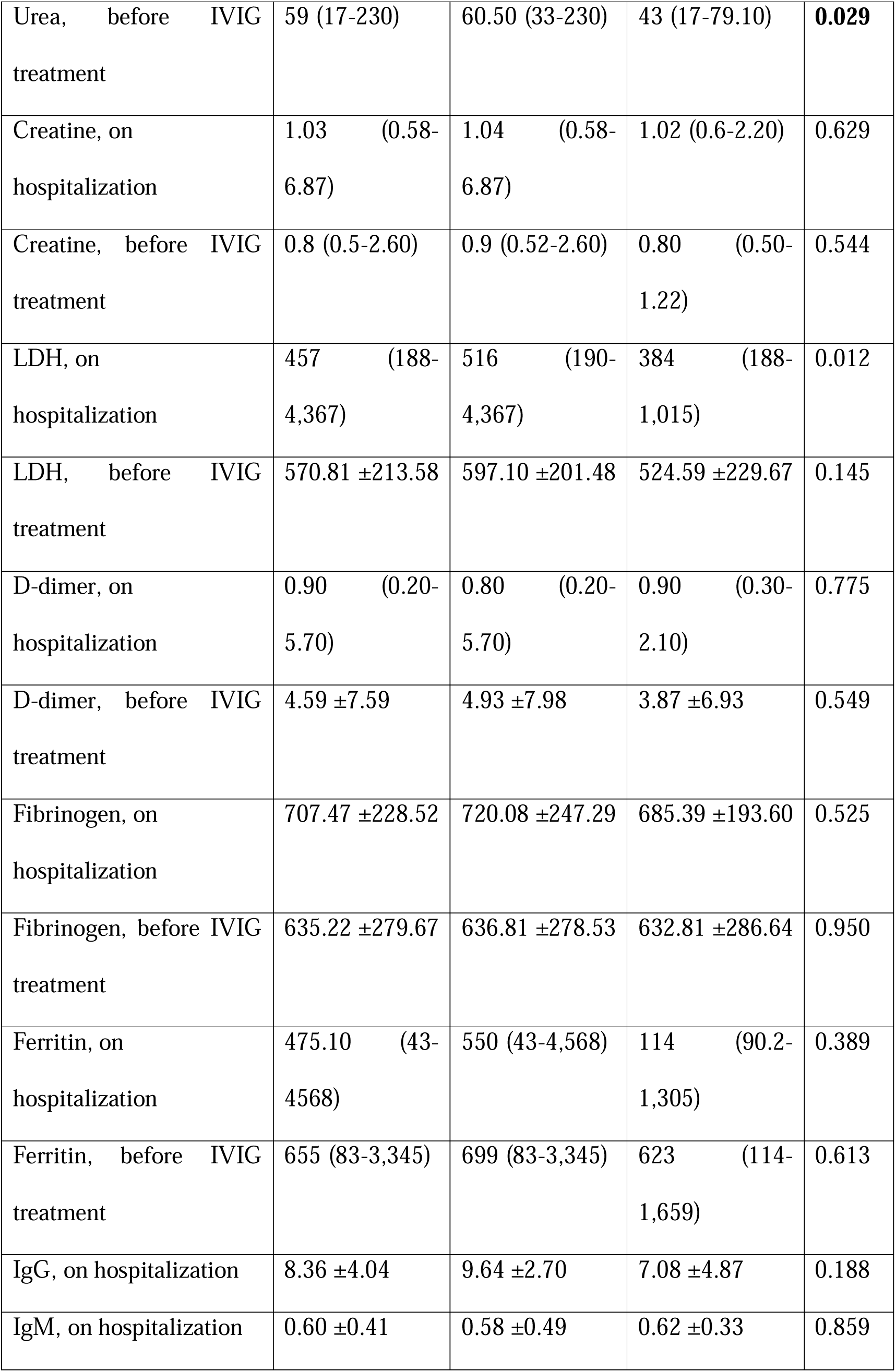
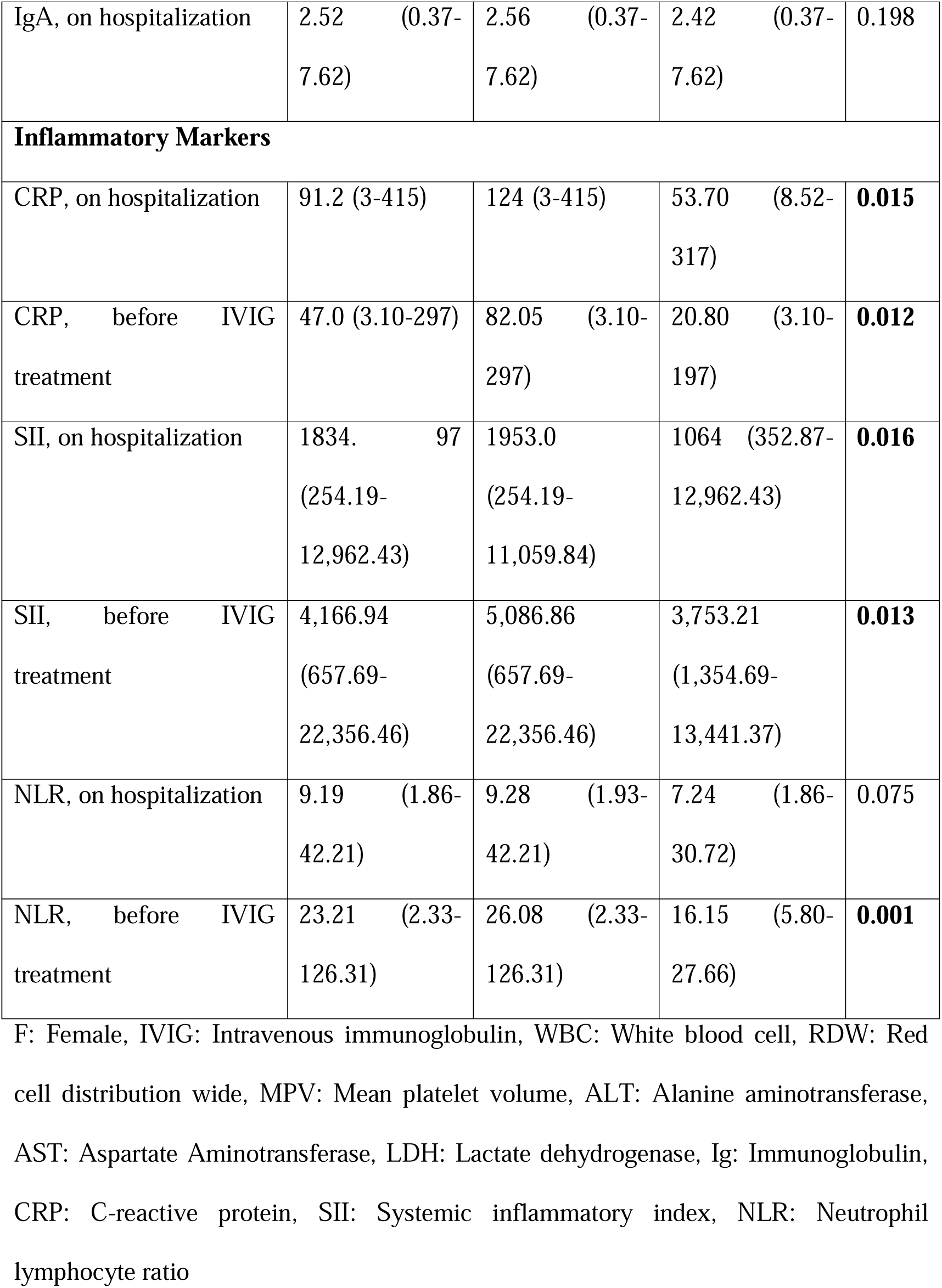
Baseline demographic, clinical, and laboratory parameters of the study population

There was no statistically significant difference between the patients who died during their follow-up and the patients who survived in terms of age, gender, tocilizumab treatment, conventional plasma treatment, and the number of days of hospitalization in the service. We observed significant differences in terms of intubated patient ratio, pulse steroid therapy, hospitalization white blood cell count, hospitalization platelet count, lymphocyte percentage before IVIG treatment, neutrophil count before IVIG treatment, hospitalization C-reactive protein (CRP) values, CRP levels before IVIG treatment, urea values before IVIG treatment, hospitalization lactate dehydrogenase (LDH) levels, hospitalization systemic inflammatory index (SII) levels, SII levels before IVIG treatment, and NLR (Neutrophil Lymphocyte Ratio) levels before IVIG treatment. The comparison of demographic, laboratory, and clinical characteristics of the patients who died and survived has been summarized in Table 1.

The univariate analysis found that the number of days of hospitalization in service, being intubated, number of IVIG treatment days, and the urea value before IVIG treatment were independent risk factors for mortality (p:0.043, p:0.001, p:0.074, p:0.004, respectively) (Table 2). As a result of multivariate analysis, being intubated and urea value before IVIG treatment were found to be independent risk factors for mortality (p:0.001 and p:0.009, respectively) (Table 3).

**Table 2:** Univariate Cox regression analyses demonstrating the relationship between baseline characteristics and Sars-CoV-2

| Variable | Univariate Analysis |  |
| --- | --- | --- |
|  | HR (95% CI) | P value |
| Day of stay in inpatient service | 0.951 (0.906 – 0.998) | **0.043** |
| Intubation | 16.370 (4.353-61.565) | **0.001** |
| Intensive care | 0.923 (0.316-0.270) | 0.884 |
| Pulse steroid therapy | 0.910 (0.525 – 1.580) | 0.738 |
| Tocilizumab treatment | 1.436 (0.794 – 2.552) | 0.236 |
| Duration of IVIG treatment, day | 1.419 (0.966 – 2.083) | **0.074** |
| IgG, on hospitalization | 1.234 (0.964 – 1.581) | 0.096 |
| WBC, before IVIG treatment | 1.000 (1.000 – 1.000) | 0.418 |
| Neutrophil count, on hospitalization | 1.000 (1.000 – 1.000) | 0.946 |
| Platelet count, on hospitalization | 1.001 (0.998 – 1.004) | 0.405 |
| Eosinophil count, IVIG öncesi | 0.994 (0.983 – 1.006) | 0.307 |
| FBG, on hospitalization | 1.002 (0.998 – 1.005) | 0.299 |
| FBG, before IVIG treatment | 1.002 (0.999 – 1.006) | 0.192 |
| Urea, before IVIG treatment | 1.011 (1.003 – 1.018) | **0.004** |
| LDH, on hospitalization | 1.000 (0.999 – 1.000) | 0.722 |
| LDH, before IVIG treatment | 1.000 (0.999 – 1.001) | 0.695 |
| IgA, on hospitalization | 1.025 (0.740 – 1.421) | 0.880 |
| CRP, on hospitalization | 1.002 (0.999 – 1.005) | 0.111 |
| CRP, before IVIG treatment | 1.001 (0.998 – 1.005) | 0.364 |
| SII, on hospitalization | 1.000 (1.000 – 1.000) | 0.746 |
| SII, before IVIG treatment | 1.000 (1.000 – 1.000) | 0.314 |
| NLR, on hospitalization | 1.002 (0.979 – 1.025) | 0.884 |
| NLR, before IVIG treatment | 1.004 (0.994 – 1.014) | 0.414 |

Sars-CoV-2: Severe Acute Respiratory Syndrome causing Coronavirus, F: Female, IVIG: Intravenous immunoglobulin, WBC: White blood cell, RDW: Red cell distribution wide, MPV: Mean platelet volume, ALT: Alanine aminotransferase, AST: Aspartate Aminotransferase, FBG: Fasting blood glucose, LDH: Lactate dehydrogenase, Ig: Immunoglobulin, CRP: C-reactive protein, SII: Systemic inflammatory index, NLR: Neutrophil lymphocyte ratio

**Table 3:** Multivariate Cox regression analyses demonstrating the relationship between baseline characteristics and Sars-CoV-2

| Variables | Multivariate Analysis |  |
| --- | --- | --- |
|  | HR (95% CI) | P value |
| Intubation | 0.389 (0.218 – 0.693) | **0.001** |
| Day of stay in inpatient service | 0.968 (0.915 – 1.024) | 0.256 |
| Duration of IVIG treatment, day | 0.833 (0.652 – 1.065) | 0.145 |
| Urea, before IVIG treatment | 1.009 (1.002 – 1.017) | **0.009** |

Sars-CoV-2: Severe Acute Respiratory Syndrome causing Coronavirus, IVIG: Intravenous immunoglobulin

It was found that for 60 mg/dL level of urea value before IVIG treatment, the sensitivity value for mortality in COVID-19 patients receiving IVIG treatment was 46.2%, and the specificity was 35.5% (p:0.029) (Table 4) (Figure 1).

**Table 4:** Sensitivity and specificity of urea level before IVIG treatment

| Risk factor | AUC (95%) | Cut-off | p | Sensitivity | Specificity |
| --- | --- | --- | --- | --- | --- |
| Urea, before IVIG treatment | 0.647<br>(0.518 - 0.776) | 60 | 0.029 | 46.2 | 35.5 |

AUC: Area under the curve, IVIG: Intravenous immunoglobulin

**Figure 1:**
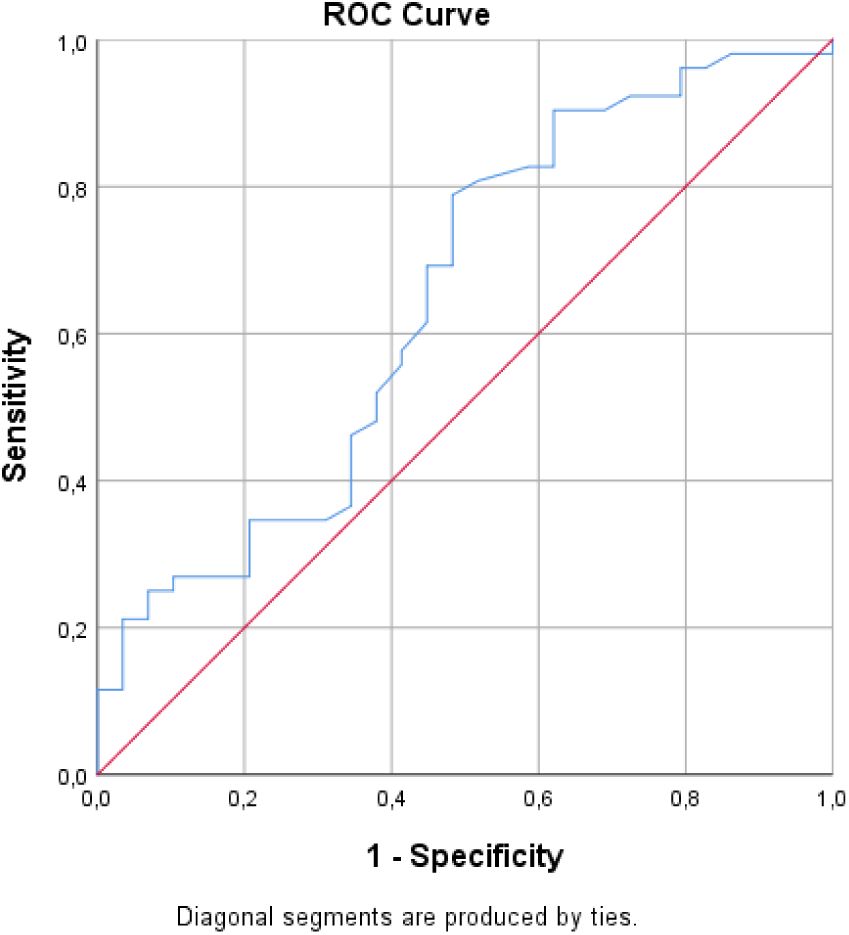
Sensitivity and specificity of urea level before IVIG treatment

## 4. Discussion

The SARS-CoV-2 virus has caused one of the most severe pandemics in human history and has put a lot of pressure, particularly on healthcare systems, since March 2020, when it was declared as a pandemic by WHO. The SARS-CoV-2 virus has caused the deaths of approximately 3 million people in nearly one year since its outbreak (2, 11). At present, there is no globally accepted treatment scheme for treating patients hospitalized for COVID-19. Therefore, it is crucial to determine the prognostic factors in vulnerable patient groups and to develop treatment modalities specific to patient groups according to these factors to reduce mortality and morbidity. In line with this opinion, this study found that being intubated and urea values before IVIG treatment are independent risk factors for COVID-19-related mortality in patients hospitalized for COVID-19 and given IVIG treatment.

It has been reported that 7% of COVID-19 patients develop acute renal failure (12, 13). In addition, renal failure has been reported to increase COVID-19-related hospital deaths in mortality studies (14-19). Cheng et al. (15) showed an increase in blood urea nitrogen increased mortality 3.97 times in COVID-19 patients. Another study reported that hospitalization blood urea nitrogen (BUN) and D-dimer levels were associated with mortality, and BUN values of ≥4.6 mmol/L included a high risk for hospital deaths (14). In another study, 6.29% of COVID-19 patients showed an increase in BUN, and increased basal BUN and creatinine values were reported to cause high mortality (17). Ng et al. (18) reported that being intubated and BUN values are risks for hospital mortality in patients with end-stage renal disease and COVID-19. Although the increase in BUN after SARS-CoV-2 is frequent, the reason for this increase is not clear. Renal epithelial cells contain angiotensin-converting enzyme 2 (ACE2) receptors that are 100 times more intense than respiratory epithelial cells; SARS-CoV-2 is internalized to renal cells and may cause renal function loss with cytopathic effect (15, 20). It has been suggested that this may increase the absorption of BUN from the renal tubules by activating the renin-angiotensin-aldosterone system (20). Although IVIG treatment is often used as one of the last treatment options in patients who do not respond to other treatments, IVIG treatment itself may be associated with renal damage (13).

On the other hand, the increase in BUN levels in COVID-19 patients may be an indicator of kidney dysfunction and an increased inflammatory state. The renal load caused by increased catabolism, hypovolemia-induced renal hypoperfusion, sepsis, drugs used in the treatment of COVID-19 such as steroid therapy, and rhabdomyolysis may also cause an increase in BUN. Although creatinine, another indicator of renal damage, was not found to be a predictor of mortality in this study, the fact that BUN is predictive of mortality suggests that BUN increases due to inflammatory conditions rather than a renal-induced reason and that increased inflammatory processes play a role in making BUN a risk factor for mortality. Another situation supporting this hypothesis is that inflammatory markers of the patients who died before IVIG treatment were prominently higher and statistically significant than the alive patients. As the most common cause of mortality in COVID-19 is a respiratory failure caused by cytokine storm, the majority of patients (81.5%) in the present study had to be followed up in the intensive care unit due to deterioration in their clinical condition. IVIG treatment is one of the last options in COVID-19 patients who are unresponsive to other therapies and whose cytokine storms are not controlled. It was thought that these patients face an intense inflammatory process, which causes an increase in BUN.

The retrospective design, relatively small study population, lack of evaluation of other renal markers such as proteinuria and hematuria, and lack of knowledge of what happened in the post-follow-up period form the main limitations of this study.

## 5. Conclusion

In conclusion, the study found that urea values before IVIG treatment were a risk factor for mortality in patients who received IVIG treatment for COVID-19. This is important as it indicates that BUN values should be closely monitored in patients given IVIG treatment for COVID-19. It also suggests that when resources are limited and risk stratification is required in COVID-19 patients, BUN values can be helpful.

## Data Availability

All data produced in the present study are available upon reasonable request to the authors

## Acknowledgement

None

